# Limitations in next-generation sequencing-based genotyping of breast cancer polygenic risk score loci

**DOI:** 10.1101/2023.12.15.23298835

**Authors:** Alexandra Baumann, Christian Ruckert, Christoph Meier, Tim Hutschenreiter, Robert Remy, Benedikt Schnur, Marvin Döbel, Rudel Christian Nkouamedjo Fankep, Dariush Skowronek, Oliver Kutz, Norbert Arnold, Anna-Lena Katzke, Michael Forster, Anna-Lena Kobiela, Katharina Thiedig, Andreas Zimmer, Julia Ritter, Bernhard H.F. Weber, Ellen Honisch, Karl Hackmann, Bioinformatics Working Group of the German Consortium for Hereditary Breast & Ovarian Cancer, Gunnar Schmidt, Marc Sturm, Corinna Ernst

## Abstract

Considering polygenic risk scores (PRSs) in individual risk prediction is increasingly becoming the standard in genetic testing for hereditary breast cancer (BC). To calculate individual BC risks, the Breast and Ovarian Analysis of Disease Incidence and Carrier Estimation Algorithm (BOADICEA) with inclusion of the BCAC 313 or the BRIDGES 306 BC PRS is commonly used. Meaningful incorporation of PRSs relies on reproducing the allele frequencies (AFs), and hence, the distribution of PRS values, expected by the algorithm. Here, the 324 loci of the BCAC 313 and the BRIDGES 306 BC PRS were examined in population-specific database gnomAD and in real-world data sets of five centers of the German Consortium for Hereditary Breast and Ovarian Cancer (GC-HBOC), to determine whether these expected AFs are achieved with next-generation sequencing-based genotyping. Four PRS loci were non-existent in gnomAD v3.1.2 non-Finnish Europeans, further 24 loci showed noticeably deviating AFs. In real-world data, between 16 and up to 22 loci were reported with noticeably deviating AFs, and were shown to have effects on final risk prediction. Deviations depended on sequencing approach, variant caller and calling mode (forced versus unforced) employed. Therefore, this study demonstrates the necessity to apply quality assurance not only in terms of sequencing coverage but also observed AFs in a sufficiently large sample, when implementing PRSs in a routine diagnostic setting. Furthermore, future PRS design should be guided by reproducibility of expected AFs in addition to the observed effect sizes.

## Introduction

The German Consortium for Hereditary Breast and Ovarian Cancer (GC-HBOC) is a consortium of interdisciplinary university centers specialized in providing counseling, genetic testing and healthcare for individuals at risk for familial breast and ovarian cancer (BC/OC). Clinical management of women found to be at increased risk for BC/OC, due to inherited pathogenic variants in established BC/OC risk genes or a strong family history of cancer, demands for accurate and age-dependent risk estimates. Numerous studies demonstrated that the effects of BC susceptibility loci, i.e., common single nucleotide variants (SNVs) and short indels, which individually contribute only slightly to individual BC risks, but whose effects can be summed up to polygenic risk scores (PRSs) which can achieve a clinically relevant degree of BC risk discrimination [1–3]. As the contribution of the PRS to BC risks has also been confirmed for carriers of a pathogenic variant in moderate-to high-penetrant BC risk genes [4–7], inclusion of PRSs in individual BC risk prediction is increasingly becoming standard in GC-HBOC centers [8].

The Breast and Ovarian Analysis of Disease Incidence and Carrier Estimation Algorithm (BOADICEA), which is implemented in the CE-marked CanRisk web interface, provides (since v5) the straightforward inclusion of genetic germline test results, cancer family history, non-genetic risk factors and PRSs in a comprehensive model [9, 10]. It is therefore widely applied for individual BC risk prediction in routine diagnostics of the GC-HBOC centers. The CanRisk web interface allows the specification of individual PRSs either as manual input (including specification of the square root of the proportion of the overall polygenic variance explained) or, for a given set of PRSs, via upload of a VCF file with the genotype or dosage information per locus to consider. Whichever method is chosen, genotyping is the responsibility of the user. For PRSs for which VCF upload is supported, CanRisk provides specifications for incorporated loci, each including the variant (chromosome, genomic position for hg19, reference and effect allele), log odds ratio (i.e., effect size) and expected AF [11]. The given alleles and AFs arise from high-throughput genotyping using one of two arrays, iCOGS13 or OncoArray [2].

In the GC-HBOC centers, the BCAC 313 BC PRS, and its modified version, the BRIDGES 306 BC PRS [12], are the preferred PRS variant sets employed for BC risk prediction. The genetic germline testing and genotyping of PRS loci is based on next-generation sequencing (NGS), e.g., using the TruRisk™ or further specifically adapted multigene panels, whole-exome or whole-genome sequencing (WGS). The BRIDGES 306 BC PRS excludes loci of the original BCAC 313 BC PRS that were found not to be appropriately designable using NGS, some of which were replaced by corresponding loci in linkage disequilibrium [12]. The assessment of designability was mainly based on sufficient read coverage for diagnostic purposes when using a multigene panel approach and mapping to human reference hg19. With the implementation of BC PRS analysis in routine diagnostics and the establishment of corresponding bioinformatic workflows, further technical challenges besides insufficient coverage were identified, e.g., missing variant calls or variant calling resulted in deviating alleles. Studies systematically assessing and comparing quality and pitfalls of germline genotyping using either arrays or NGS approaches, are rare and mainly date from the early days of the establishment of NGS in clinical diagnostics [13–16]. Hence, it cannot be excluded that the conclusions drawn (which were also contradictory with regard to NGS or array being the more reliable and preferable approach) were based on now predominantly outdated technologies. Nevertheless, it is well-known that accuracy of NGS tend to be hampered in genomic regions of low complexity, i.e., homopolymer runs, tandem repeats and strongly biased GC contents, among others [17–19]. In the Genome Aggregation Database (gnomAD), the largest and most widely used population-specific variant database, variants located in so-called low-complexity regions are flagged, to indicate that reported AFs may be erroneous [20, 21].

In this study, the Bioinformatics Working Group of the GC-HBOC conducted a systematic evaluation across GC-HBOC centers to develop a detailed, locus-wise assessment of technical pitfalls and possible sources of error in NGS-based PRS genotyping. A three-stage approach was followed. First, the AF of PRS variants were compared to the gnomAD AF for the European general population and it was checked if the variants can be converted to the hg38 reference genome. Second, PRS variant AFs in real-world data sets provided by participating GC-HBOC centers were compared to the AFs expected by CanRisk. Third, possible workarounds for use in clinical diagnostics, i.e., usage of alternative alleles and proxys, were identified. The presented results are of relevance beyond diagnostics for BC risk prediction, as they demonstrate principle difficulties in NGS-based PRS computation, especially for PRSs developed based on array data. Furthermore, the results underline the necessity of a comprehensive technical evaluation of PRS variant genotyping in clinical use, as the predictive ability of an individual PRS crucially depends on the assumptions made about the underlying AFs.

## Materials and Methods

### Evaluation of expected allele frequencies & convertibility to hg38

Two BC PRS variant sets were considered, namely of the BCAC 313 and the BRIDGES 306 BC PRS. Of the two sets, 295 loci are identical, 18 loci are unique to BCAC 313 BC PRS, and further 11 loci are unique to the BRIDGES 306 BC PRS, resulting in a total number of N = 324 variants to be considered. Expected AFs were extracted from the corresponding PRS specification files at the CanRisk knowledge base [11]. Additionally, AFs for non-Finnish Europeans (NFEs) were obtained from the gnomAD v3.1.2 database^1^, which are based on more than 33,000 WGS samples mapped to the hg38 reference sequence. For conversion of the hg19-based PRS variants from CanRisk to hg38, the gnomAD liftover feature was used.

Besides AFs, gnomAD flags and warnings indicating possible technical artifacts were retrieved and recorded. These included localization within low-complexity regions, low-quality sites (i.e., sites that are covered in less than 50% of considered samples [20]) and sites not passing the allele-specific GATK Variant Quality Score Recalibration (VQSR) filter.

### Determination of deviating allele frequencies

To determine PRS variants with considerably deviating AFs, thresholds had to be defined dependent on sample sizes and variances observed. Therefore, individual thresholds per data set were determined, using an elbow of the curve method. The absolute differences between observed and expected AFs were sorted in descending order, and the absolute difference referring to the point with the largest Euclidean distance to the imaginary line between thought points (0, 1) and (N + 1, 0), were chosen as threshold, i.e., all observed absolute differences greater than this threshold were determined as noticeably deviating. Corresponding curves are shown in Supplementary Figures 1 to 6. If the same set of samples was processed with two different variant callers, the smaller threshold was applied in each case, to facilitate comparing variant caller performance.

### Real-world data collection

Genotyping results for either BCAC 313 or BRIDGES 306 BC PRS loci in a sample of at least 100 individuals of European ancestry were requested from GC-HBOC centers. Participating centers submitted observed AFs per locus as well as fractions of samples that did not meet required quality criteria (e.g., with regard to minimum read depth). Furthermore, details on sequencing approaches and bioinformatic analysis workflows for PRS genotyping were systematically recorded.

In total, five GC-HBOC centers provided data, namely the Institute of Medical Genetics and Applied Genomics (IMGAG), University Hospital Tübingen, the Institute for Clinical Genetics (ICG), University Hospital Carl Gustav Carus Dresden, the Institute of Human Genetics at the University of Münster (IHG-M), the Center for Familial Breast and Ovarian Cancer (CFBOC), University Hospital Cologne, and the Institute of Human Genetics at the University of Regensburg (IHG-R). Each center provided two NGS-based data sets. An overview on data characteristics is given in Table 1. A more detailed description of sample compositions, sequencing approaches and bioinformatic analyses can be found in Supplementary Methods.

**Table 1:**
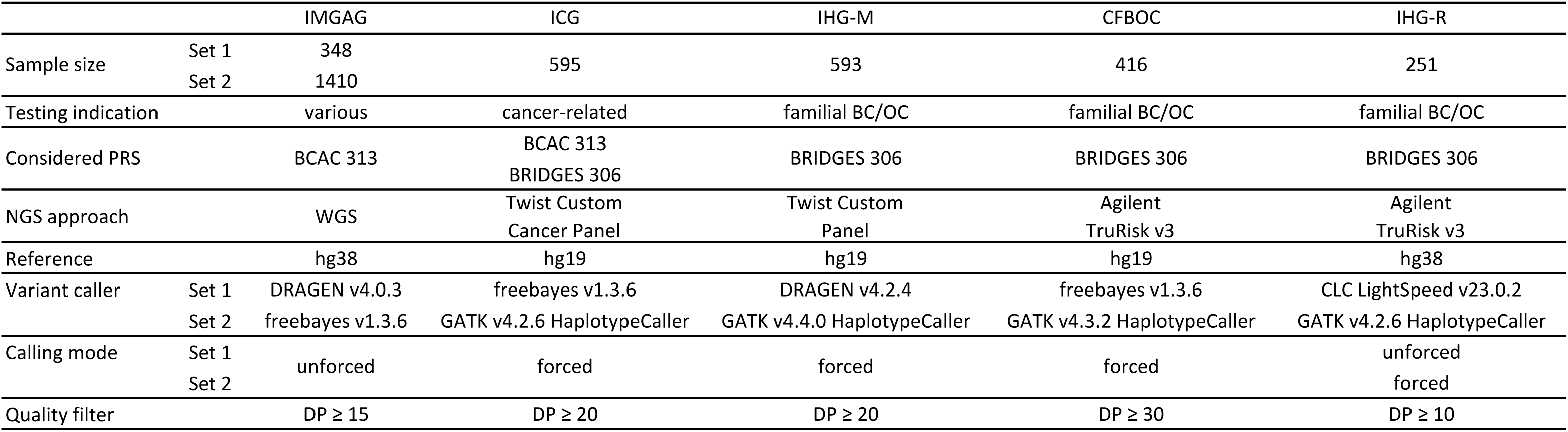
Characteristics of data sets provided by participating centers of the German Consortium for Hereditary Breast & Ovarian Cancer (GC-HBOC), namely the Institute of Medical Genetics and Applied Genomics (IMGAG), University Hospital Tübingen, the Institute for Clinical Genetics (ICG), University Hospital Carl Gustav Carus Dresden, the Institute of Human Genetics at the University of Münster (IHG-M), the Center for Familial Breast and Ovarian Cancer (CFBOC), University Hospital Cologne, and the Institute of Human Genetics at the University of Regensburg (IHG-R). Each center provided two data sets. BC/OC: Breast/ovarian cancer; DP: Sequencing depth; PRS: Polygenic risk score.

### Assessment of effects of deviating allele frequencies on estimated breast cancer risks

Effects of noticeably deviating AFs of PRS loci on CanRisk-based estimated BC risks, rely on a multitude of factors, such as the number and combination of affected loci, and additional risk factors such as results of germline testing of established BC/OC risk genes, BC/OC family history, non-genetic risk factors and current age. In principle, the effect of the PRS on BC risk is expected to be decreased in carriers of a pathogenic germline variant in a BC risk gene with moderate or high penetrance, and furthermore, its effect is expected to decrease with age [10]. In order to get an estimate of expected biases in predicted BC risks due to potentially erroneous PRS genotyping, estimates of 10 year and remaining lifetime risks, i.e., cumulative risks of primary BC until age of 80 years, were calculated using the CanRisk web interface for imaginary cancer-unaffected women of three different ages, namely 20, 40, and 60 years, without any further information than (artificial) PRS.

To simulate different scenarios, artificial VCF files were constructed with an average PRS (50th percentile) by using two times the expected CanRisk AF, i.e., expected dosage. For each data set, for loci showing noticeably deviating AFs, expected dosages were replaced by two times the observed AF in the data set. Dates of birth were set to January 1 in 2003, 1983, and 1963, to simulate 20, 40, and 60 years of age at time of risk computation, which were performed in October 2023, using CanRisk v2.3.5.

### Elaboration of workarounds

Potential solutions for improving genotyping performance with respect to expected AFs could be (besides improving the calling itself) the consideration of alternative alleles or proxys. Details on the identification of potential variants to substitute for this purpose are given in Supplementary Methods. Alternative variants in gnomAD v.3.1.2 with an AF matching the expected CanRisk AF, were further evaluated using the IMGAG freebayes data, as this (i) was the largest data set in the study (n=1410), and (ii) the only WGS-based data set, which allowed genotyping of the entire set of putative proxys.

## Results

### Missing loci & convertibility to hg38

For four BC PRS loci, no variants were listed at the specified genomic position in gnomAD v2.1.1, namely rs572022984, rs113778879, rs73754909, and rs79461387. gnomAD v3.1.2 also reported no variants for three of these four loci for corresponding loci in hg38 as defined by dbSNP [22] (Supplementary Table 1). Locus rs572022984 was listed, but with an overall allele count of zero in NFE samples (Table 2).

**Table 2:**
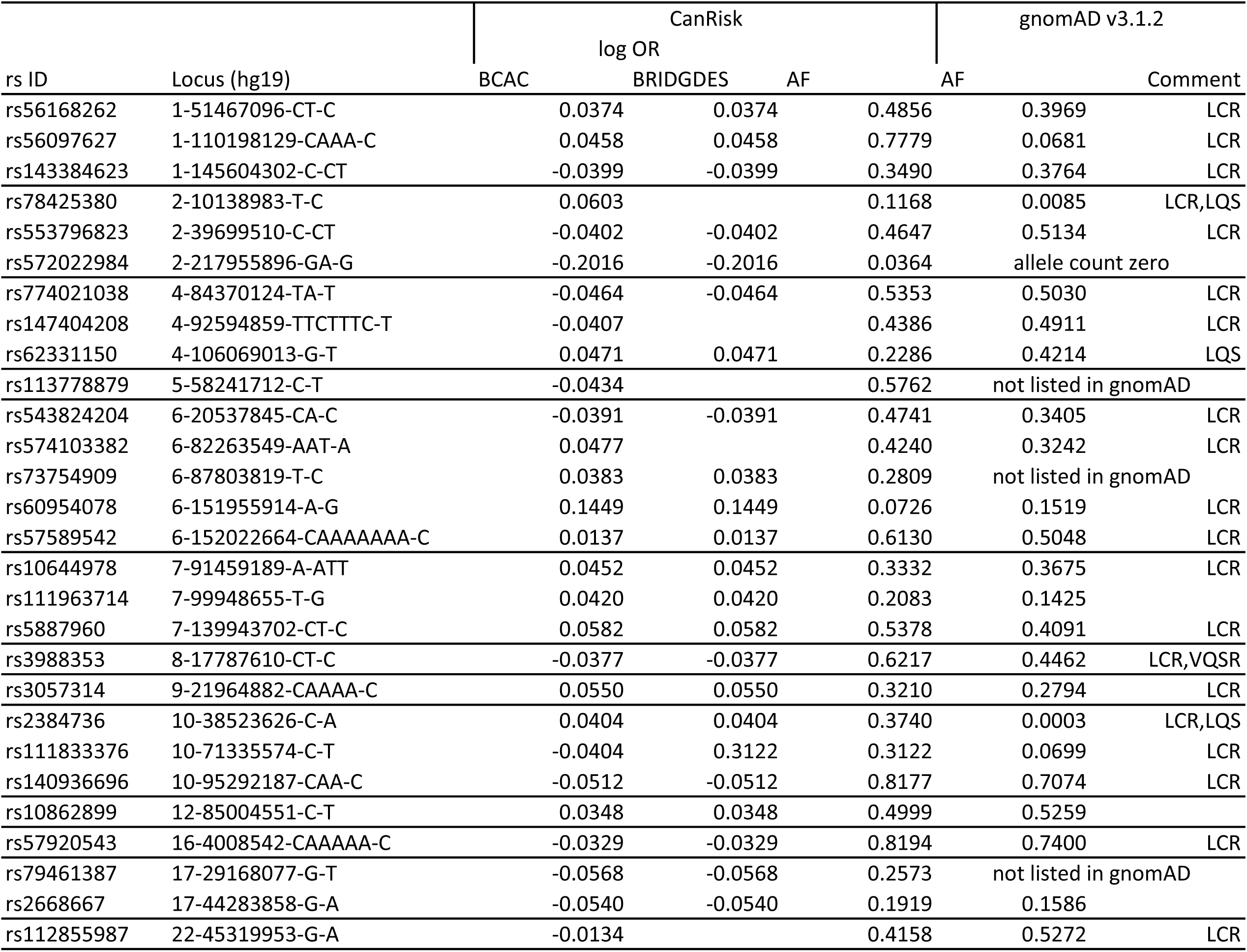
Characteristics of loci incorporated in the BCAC 313 or BRIDGES 306 breast cancer PRSs that were either not included in the gnomAD v3.1.2 database or reported with extremely deviating allele frequency compared to CanRisk. Log odds ratios (ORs) are identical for BCAC 313 and BRIDGES 306, but missing values indicate loci not included in the corresponding PRS. Entries in the Comment column refer to technical artifacts reported in gnomAD. LCR: low complexity region; LQS: low-quality site (in <50% of samples covered); VQSR: failed allele-specific GATK Variant Quality Score Recalibration (VQSR) filter.

For two loci, conversion to hg38 resulted in a change in alleles, namely for rs143384623 (hg19: 1-145604302-C-CT; hg38: 1-145830798-C-CA) and rs550057 (hg19: 9-136146597-C-T; hg38: 9-133271182-T-C). For rs143384623, the change of the alternative allele from CT to CA did not result in a noticeable shift in AFs observed in gnomAD NFE samples (5142/13304 (0.39) in v2.1.1 versus 24316/64610 (0.38) in v3.1.2, two-sided Fisher’s exact test p = 0.14). For rs550057, the observed AFs appeared exactly opposite, i.e., 3786/14828 (0.26) for allele T in gnomAD v2.1.1 and 49878/67552 (0.74) for allele C in gnomAD v3.1.2. Therefore, 1 − 49878/67552 was assumed as the gnomAD v3.1.2 effect AF at this bi-allelic site.

### Allele frequencies & technical artifacts reported in gnomAD v3.1.2

For 39 of the 320 PRS loci listed with AF*>*0 in gnomAD v3.1.2, at least one observation of technical artifacts was reported: 38 loci were flagged as being located in low complexity regions, three as being localized at a low-quality site, and one failed the allele-specific VQSR filter (Supplementary Table 1).

Due to the absolute difference threshold 0.016 (Supplementary Figure 1), 24 loci were determined as showing deviating AFs compared to CanRisk (Figure 1, Table 2). Absolute differences ranged from 0.03 to 0.71, and for 21 out of these 24 loci (87.5%), technical artifacts were reported in gnomAD v3.1.2.

**Figure 1:**
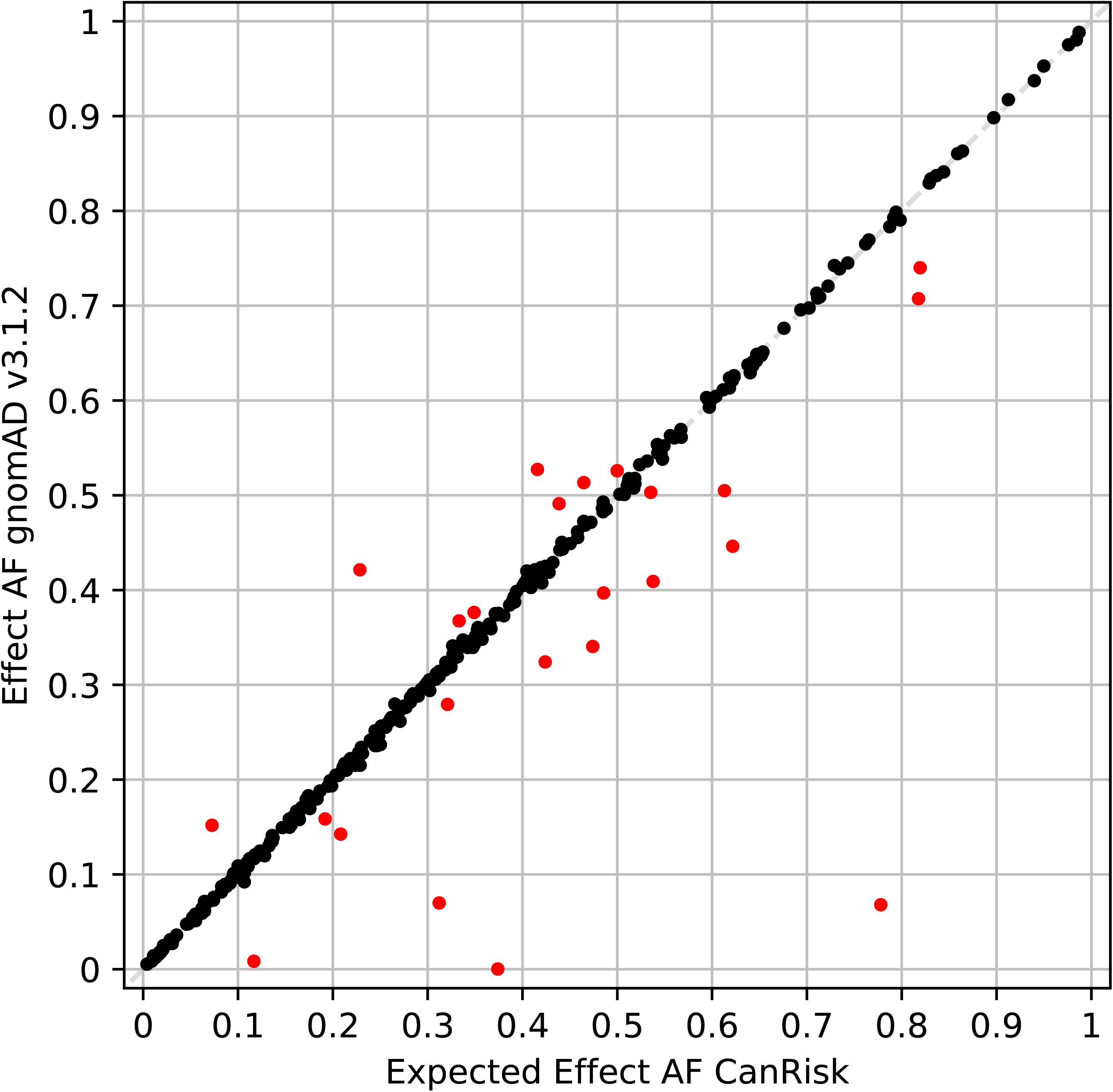
Comparison of variant effect allele frequencies (AFs) specified by CanRisk and observed in gnomAD v3.1.2 non-Finnish European samples for 320 variants incorporated in BCAC 313 or BRIDGES 306 breast cancer polygenic risk scores. Extremely deviating AFs with absolute difference > 0.016 are indicated by red markers.

### Evaluation of real-world next-generation sequencing outcome

All 50 PRS loci for which a noticeably deviating AF was observed in at least one of the data sets provided by the five participating GC-HBOC centers are listed in Table 3.

**Table 3:**
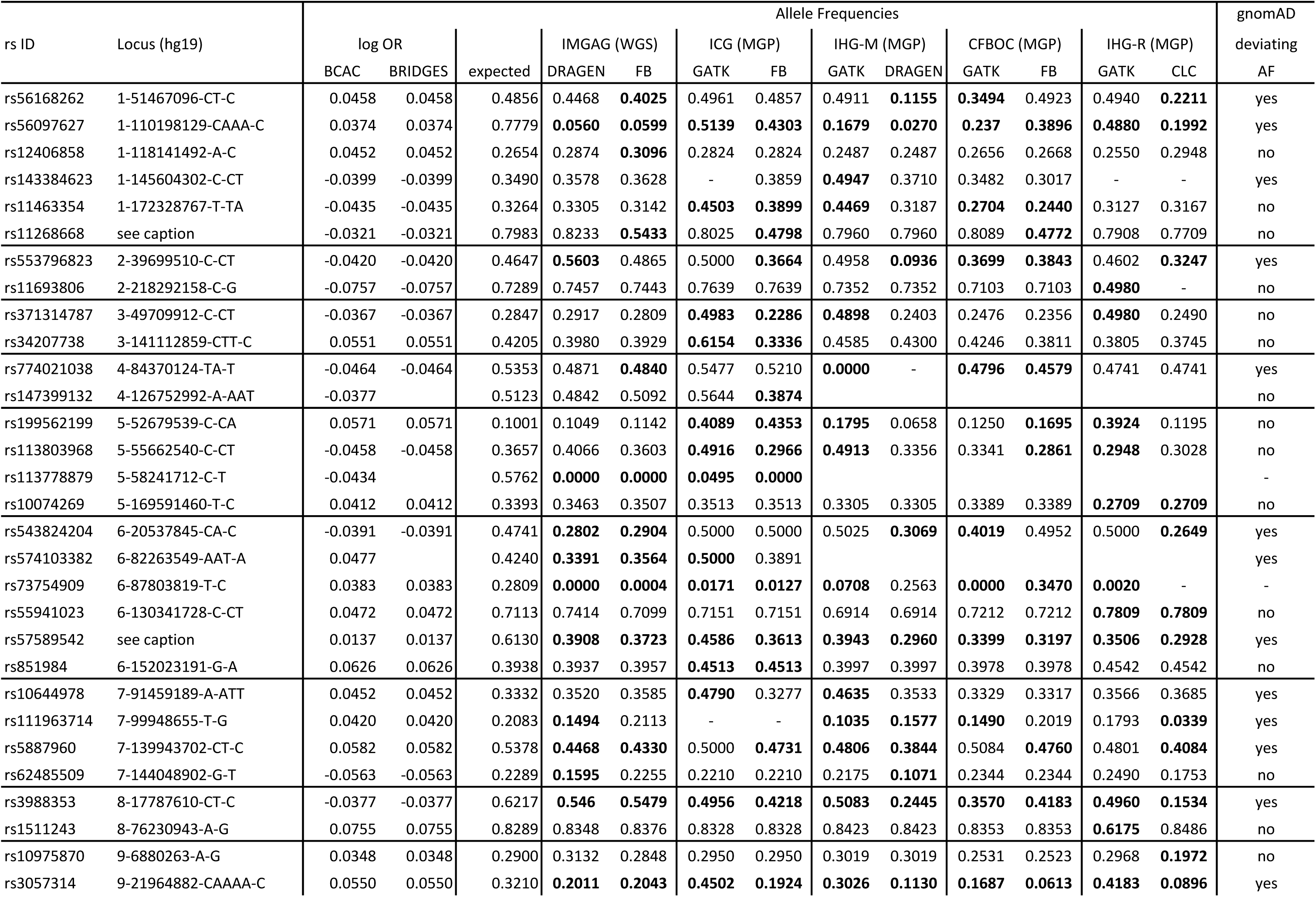
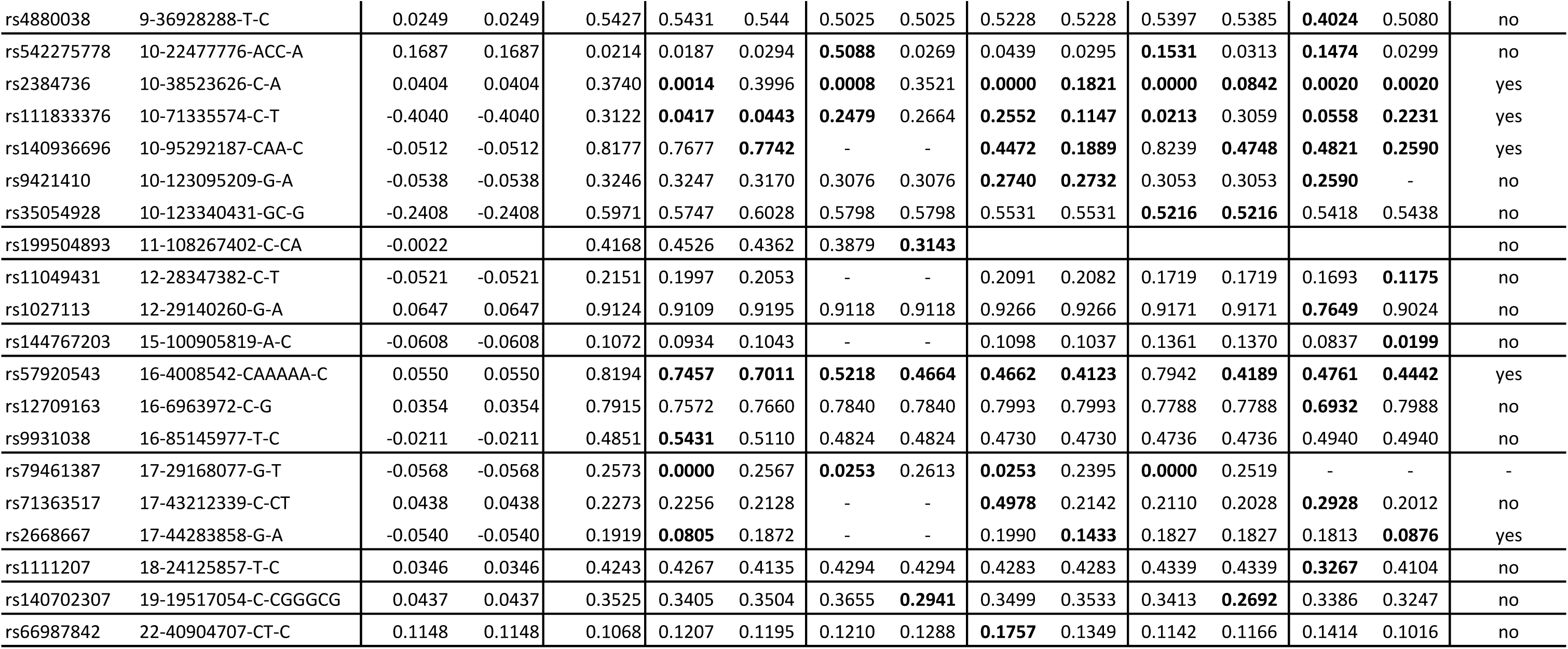
Summary of polygenic risk score genotyping results with noticeably deviating allele frequencies (AFs) of centers of the German Consortium for Hereditary Breast and Ovarian Cancer. Noticeably deviating AFs are shown in bold. Loci (hg19-based) of rs11268668 and rs57589542 are 1-204502514-T-TTCTGAAACAGGG (hg19) and 6-152022664-CAAAAAAA-C (hg19), respectively. WGS: Whole-genome sequencing. MGP: Multi-gene panel sequencing. FB: freebayes.

For the IMGAG DRAGEN data, 0.052 was calculated as threshold to determine noticeably deviating AFs (Supplementary Figure 2), resulting in 18 loci affected (Table 3, Figure 2). Of these, 16 were previously also identified as missing or showing noticeably deviating AFs in gnomAD v3.1.2. The exceptions were rs62485509 and rs9931038. For IMGAG freebayes data, 0.036 was calculated as threshold (Supplementary Figure 2), resulting in 16 loci from the BCAC 313 BC PRS determined as showing a noticeably deviating AF. Of these, 11 loci were also identified as showing deviating AF in IMGAG DRAGEN data, and all but rs12406858 and rs11268668 were previously identified as missing or showing deviating AFs in gnomAD v3.1.2.

**Figure 2:**
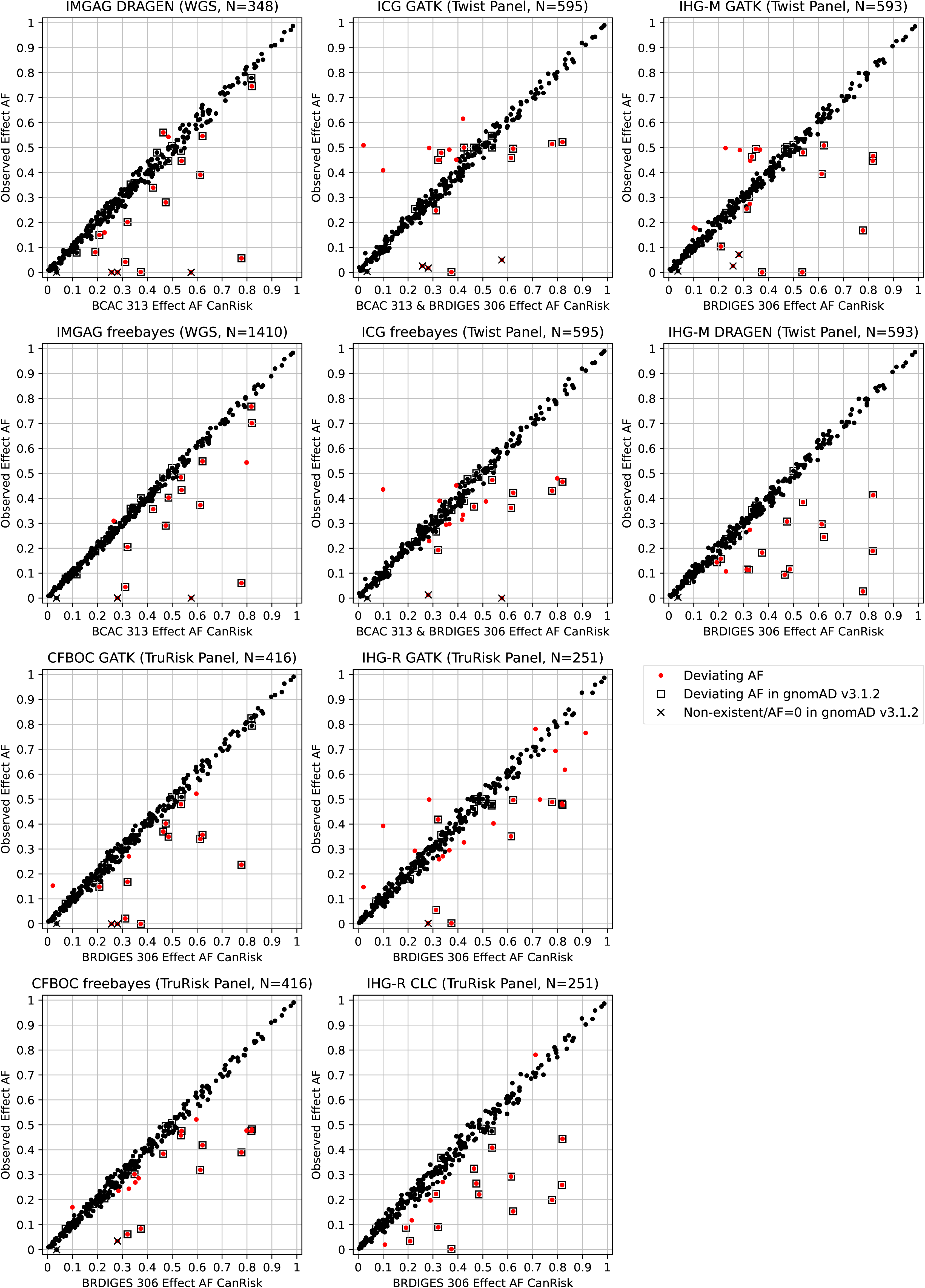
Comparison of effect allele frequencies (AFs) specified by CanRisk and observed in 10 real-world data sets for 320 loci incorporated in BCAC 313 or BRIDGES 306 breast cancer polygenic risk scores. Data was provided by the Institute of Medical Genetics and Applied Genomics (IMGAG) at University Hospital Tübingen, Institute for Clinical Genetics (ICG) at University Hospital Carl Gustav Carus Dresden, by the Institute of Human Genetics at the University of Münster (IHG-M), by the Center for Familial Breast and Ovarian Cancer (CFBOC) at University Hospital Cologne, and by the Institute of Human Genetics at the University of Regensburg (IHG-R).

Considering genotyping data provided by ICG, 23 of the overall 324 PRS loci did not meet the minimum quality criteria (read depth *≥*20) in more than 25% of samples and were discarded (Supplementary Table 2). Additionally, GATK reported read depth *<*20 for *>*25% of samples for rs143384623. For 266 of the remaining 300 PRS loci (88.67%), forced genotyping with GATK and freebayes resulted in observation of identical AFs. For both ICG GATK and freebayes data, 0.053 was calculated as threshold to determine noticeably deviating AFs (Supplementary Figure 3). Using this threshold, 19 loci showed noticeably deviating AFs in each dataset (including two loci exclusive for BCAC 313 BC PRS), with an overlap of 13 (Table 3, Figure 2).

The IHG-M provided GATK-and DRAGEN-based BRIDGES 306 BC PRS genotyping data of 593 samples. Locus rs138179519 did not meet the quality criteria, and additionally rs774021038 using DRAGEN. Of the remaining 304 loci, 252 (82.89%) showed identical AFs (Supplementary Table 2). Using a threshold of 0.046 (Supplementary Figure 4), resulted in 22 loci showing deviating AFs in GATK data, respectively 16 loci in DRAGEN data, with an overlap of 11 loci.

For the CFBOC data based on 416 samples, a threshold of 0.046 was calculated (Supplementary Figure 5). The loci of the BRIDGES 306 BC PRS were considered, 243 (79.41%) of which showed identical AFs for both callers applied (Supplementary Table 2). Overall 23 loci (all of which are included also in the BCAC 313 BC PRS) showed deviating AFs: 16 loci in GATK and 17 loci in freebayes data, with an overlap of 10 loci.

The IHG-R provided GATK-and CLC-based BRIDGES 306 BC PRS genotyping data of 251 samples (Supplementary Methods). Four loci did not meet the quality criteria in both settings, and additional four in the CLC setting. Of the remaining 298 loci, 228 (76.51%) showed identical AFs (Supplementary Table 2). Using a threshold of 0.063 (Supplementary Figure 6), resulted in 23 loci showing noticeably deviating AFs in GATK data, respectively 19 loci in CLC data, with an overlap of 10 loci.

In summary, for five loci, deviating AFs were reported in all GC-HBOC real-world settings examined, namely for rs56097627, rs113778879, rs57589542, rs3988353, and rs3057314. Further three loci, namely rs574103382, rs73754909, and rs57920543, were reported with deviating AFs in all settings except for one (Table 3).

However, there were also 13 loci that were conspicuous in a single setting exclusively, namely four in IHG-R GATK data (rs1511243, rs4880038, rs1027113, rs1111207), three in IHG-R CLC-data (rs10975870, rs11049431, rs144767203), two each in ICG freebayes data (rs147399132, rs199504893) and IHG-M GATK data (rs143384623, rs66987842), and one each in IMGAG DRAGEN (rs9931038) and IMGAG freebayes data (rs12406858). Another 6 loci (rs34207738, rs10074269, rs55941023, rs851984, rs9421410, rs35054928) showed AF deviations in only one center, but these were concordant.

Considering the loci non-existent in gnomAD v3.1.2, rs113778879 was not observed with expected AF in any GC-HBOC center, and rs73754909 only with forced DRAGEN calling in IHG-M data. For rs79461387, expected AFs were reported when using freebayes or forced DRAGEN calling only. Of note, rs572022984 with zero allele count in gnomAD v3.1.2 NFEs and an expected AF of 0.0364 in CanRisk, was consistently not observed at all or with a maximum AF of 0.005 (Supplementary Table 2).

Five loci showing aberrant AFs in gnomAD v3.1.2 NFEs (Table 2) were not reported with deviating AF by any of the participating GC-HBOC centers, namely rs78425380, rs62331150, rs60954078, rs10862899, and rs112855987.

### Implications on risk prediction

Without further information and assuming a standardized PRS at the 50th percentile, the estimated 10 year risks of developing primary BC of cancer-unaffected women of 20, 40, and 60 years of age were 0.1%, 1.5%, and 3.4% according to CanRisk (Supplementary Table 3). Percentiles of PRSs from artificial VCF files with aberrant dosages (see Methods) ranged from 47.5% (IHG-R CLC, BRIDGES 306) up to 55.3% (ICG freebayes, BCAC 313). The risk of 0.1% for a 20 year old woman was concordantly unchanged in all scenarios including artificial PRSs. For a 40 year old woman, estimated 10 year risks were increased by 0.1% in seven scenarios, and for a 60 year old woman by up to 0.2% in nine scenarios.

Estimated remaining lifetime risks of developing primary BC assuming an average PRS (50th percentile) of cancer-unaffected women aged 20, 40, and 60 years are 11.3%, 10.9%, and 7.1% according to CanRisk (Supplementary Table 3). When using PRSs from artificial VCF files with aberrant dosages, estimated lifetime risks ranged from 11.1% up to 11.9% for a 20 year old woman, from 10.6% up to 11.4% for a 40 year old woman, and from 7.0% up to 7.4% for a 60 year old woman, whereby the lowest estimates were obtained with the BRIDGES 306 BC PRS based on IHG-R CLC data with 19 artificial dosages imputed, and the highest with the BCAC 313 BC PRS based on ICG freebayes data with also 19 artificial dosages imputed.

### Consideration of alternative alleles and loci in linkage disequilibrium

For 20 PRS loci showing noticeably deviating AFs in at least one real-world NGS data set, alternative alleles or overlapping variants with minimum AF 0.01 in NFEs were reported in gnomAD v3.1.2 (Supplementary Table 4). For rs73754909 and rs79461387, both SNVs and non-existent in gnomAD v3.1.2, deletions were reported with comparable AFs to the ones expected by CanRisk. For both deletions, the adjacent downstream nucleotide of the reference sequence was identical to the substituted nucleotide of the expected effect allele (Figure 3). For rs113778879, which is also an SNV not contained in gnomAD v3.1.2, a similar observation could be made (Supplementary Figure 7), but the reported AF exceeds the expected one by more than 0.1 (0.5762 versus 0.6818).

**Figure 3:**
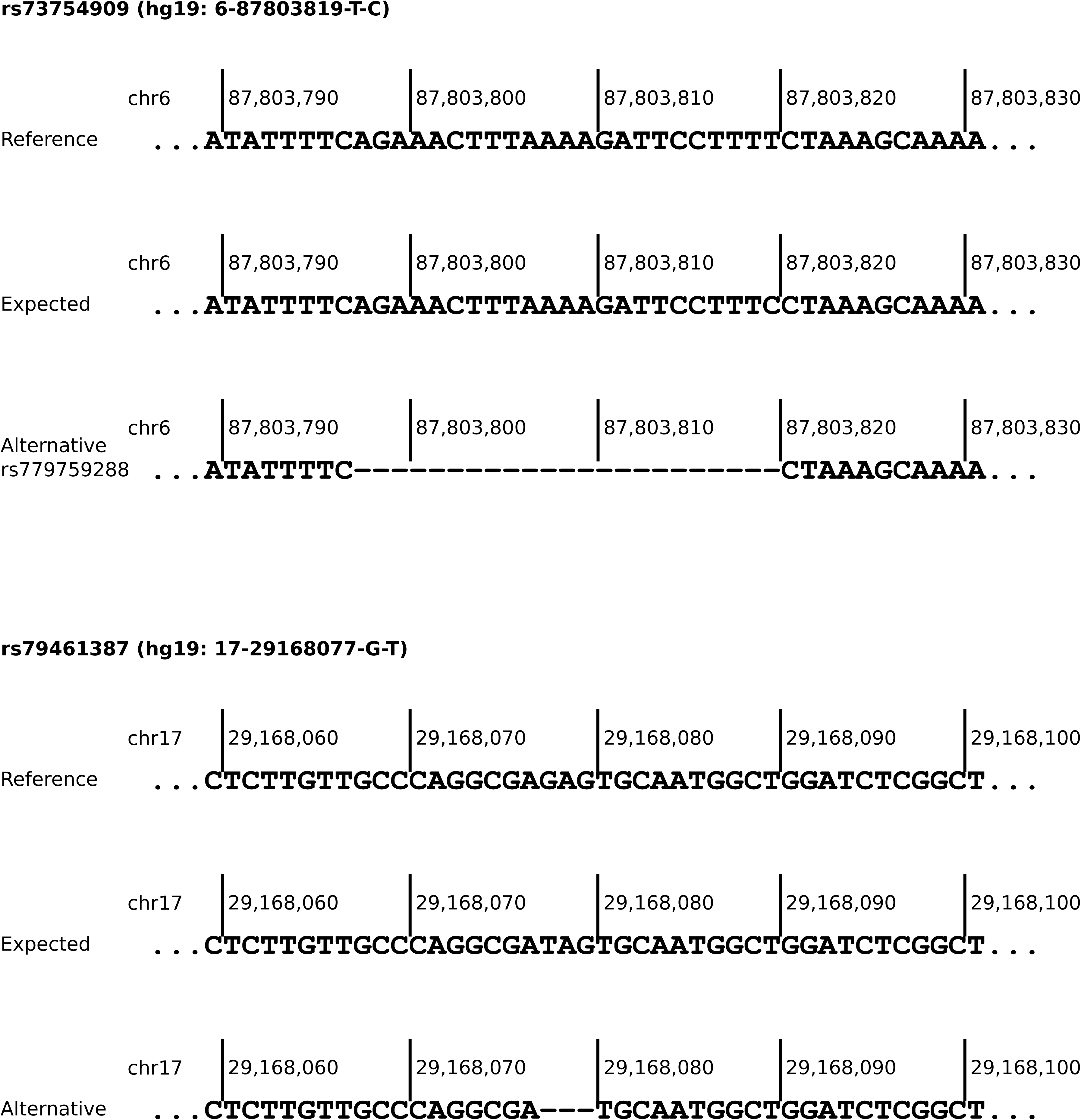
Sequences of reference, expected effect allele and potential alternative allele of polygenic risk score loci rs73754909 and rs79461387 (hg19-based). Both alternative alleles are deletions with the adjacent downstream nucleotide identical to the expected substituted one.

For 29 out of the 50 loci showing noticeable deviating AFs in at least one real-world data set, proxys in 1000G GRCh37 microarray data, 1000G GRCh38 High Coverage WGS data, or TOPMED European data could be identified (Supplementary Table 5). For rs73754909, rs79461387, and rs113778879, LDpair based on GRCh38 reported the same alternative alleles as gnomAD v3.1.2 (Supplementary Table 4), where the original PRS loci are non-existent.

Proxys and alternative alleles showing AFs in gnomAD v3.1.2 comparable to expected CanRisk AFs, i.e., an absolute deviation *<*0.016, were considered as possible workarounds for improved PRS genotyping, and further evaluated with respect to observed AFs in IMGAG freebayes data (Table 4). For 20 of these 22 PRS loci, absolute differences between expected and observed AFs in IMGAG freebayes data remained below the previously defined IMGAG freebayes-specific threshold of 0.036. The exceptions were the substitutions of rs12406858 and rs79461387. The latter is noteworthy because the original PRS locus, which is an SNV, is correctly called by freebayes in forced and unforced mode (Table 3), whereas GATK HaplotypeCaller seems to call an overlapping deletion of sequence GAG. Also noteworthy are the potential replacements of rs73754909 and rs111833376, as both variants are consistently called with noticeably deviating AFs in real-world data sets.

**Table 4:** Potential solutions for improving polygenic risk score (PRS) genotyping performance with respect to the achievement of allele frequencies (AFs) expected by CanRisk, using alternative alleles or proxys. Resulting AFs were investigated based on gnomAD v3.1.2 non-Finnish European data and genotyping results of 1410 European whole-genome sequencing (WGS) samples using (unforced) freebayes, provided by the Institute of Medical Genetics and Applied Genomics (IMGAG) at University Hospital Tübingen.

| rs ID | Locus (hg19) | Expected AF | Workaround | gnomAD AF | IMGAG FB AF |
| --- | --- | --- | --- | --- | --- |
| rs12406858 | 1-118141492-A-C | 0.2654 | Proxy rs1966228 | 0.2622 | 0.3064 |
| rs11693806 | 2-218292158-C-G | 0.7289 | Proxy rs3821098 | 0.7422 | 0.7443 |
| rs34207738 | 3-141112859-CTT-C | 0.4205 | Summing up the AFs of deletions of two and three thymines | 0.4344 | 0.4167 |
| rs10074269 | 5-169591460-T-C | 0.3393 | Proxy rs4562056 | 0.3414 | 0.3511 |
| rs73754909 | 6-87803819-T-C | 0.2809 | Alternative allele rs77846138* | 0.2846 | 0.2578 |
|  |  |  | Proxy rs12664322 | 0.2849 | 0.2791 |
| rs55941023 | 6-130341728-C-CT | 0.7113 | Proxy rs1415700 | 0.7049 | 0.7050 |
|  |  |  | Proxy rs11390217 | 0.7058 | 0.7046 |
| rs851984 | 6-152023191-G-A | 0.3938 | Proxy rs851983 | 0.3993 | 0.3965 |
| rs1511243 | 8-76230943-A-G | 0.8289 | Proxy rs6472903 | 0.8294 | 0.8376 |
| rs10975870 | 9-6880263-A-G | 0.29 | Proxy rs12380608 | 0.2863 | 0.2840 |
|  |  |  | Proxy rs10975887 | 0.2849 | 0.2837 |
| rs4880038 | 9-36928288-T-C | 0.5427 | Proxy rs4880039 | 0.5449 | 0.5436 |
|  |  |  | Proxy rs7032313 | 0.5446 | 0.5440 |
| rs542275778 | 10-22477776-ACC-A | 0.0214 | Proxy rs112287594 | 0.0185 | 0.0270 |
| rs111833376 | 10-71335574-C-T | 0.3122 | Summing up AFs of rs111833376 and rs753981427† | 0.3200 | 0.3163 |
|  |  |  | Proxy rs12769661 | 0.2984 | 0.2929 |
| rs9421410 | 10-123095209-G-A | 0.3246 | Proxy rs7913694 | 0.3142 | 0.3110 |
|  |  |  | Proxy rs35098964 | 0.3139 | 0.3099 |
| rs35054928 | 10-123340431-GC-G | 0.5971 | Proxy rs2981579 | 0.5908 | 0.5996 |
| rs11049431 | 12-28347382-C-T | 0.2151 | Proxy rs11049519 | 0.2142 | 0.2039 |
| rs144767203 | 15-100905819-A-C | 0.1072 | Proxy rs58855876 | 0.1078 | 0.1043 |
|  |  |  | Proxy rs113438754 | 0.1078 | 0.1043 |
| rs12709163 | 16-6963972-C-G | 0.7915 | Proxy rs1492386 | 0.7951 | 0.7684 |
| rs9931038 | 16-85145977-T-C | 0.4851 | Proxy rs60296580 | 0.4903 | 0.5082 |
| rs79461387 | 17-29168077-G-T | 0.2573 | Alternative allele rs550458309‡ | 0.2719 | 0.0000 |
| rs2668667 | 17-44283858-G-A | 0.1919 | Proxy rs2532237 | 0.1860 | 0.1957 |
|  |  |  | Proxy rs150290194 | 0.1765 | 0.1858 |
| rs1111207 | 18-24125857-T-C | 0.4243 | Proxy rs1111208 | 0.4249 | 0.4135 |
| rs66987842 | 22-40904707-CT-C | 0.1068 | Proxy rs6001949 | 0.1003 | 0.1195 |

\*6-87094100-CAGAACTTTAAAAGATTCCTTTT-C (hg19)
†10-71335572-TCC-T (hg19)
‡17-29168076-AGAG-A (hg19)

## Discussion

This study describes the systematic evaluation of NGS-based PRS genotyping in real-world data sets of five GC-HBOC centers. The observed AFs of PRS loci in individuals with European descent were employed as quality criterion, as the reproducibility of expected AFs of the PRS loci, and hence, the assumptions made about the overall PRS distribution, are an essential prerequisite for a correct risk calculation. In each setting under consideration, at least 14 out of 313 BCAC BC PRS loci, respectively 306 BRIDGES BC PRS loci, showed noticeably deviating AFs. These deviations were dependent on sequencing technology, variant caller and calling mode and can be expected to affect final BC risk calculations of the BOADICEA model implemented in CanRisk. Therefore, this study demonstrates the necessity to apply quality assurance not only in terms of sequencing coverage but also in terms of observed AFs in a sufficiently large cohort, when implementing PRSs in a routine diagnostic setting.

The presented results also point to potential solutions for improving genotyping performance with respect to the achievement of expected AFs for several loci, these primarily include the use of alternative variant callers or consideration of proxy variants. The use of certain variant callers resulted consistently in noticeable deviating AFs, which were not observed for other callers. This concerned e.g. rs62485509 when using DRAGEN, and rs11268668 when using freebayes (Table 3). In each setting under investigation considering identical samples, the number of loci whose AFs match the expected AFs could be increased by variant-specific selection of the variant caller.

Comparison to large-scale population-specific data, such as gnomAD and 1000G High Coverage WGS, indicates that several PRS loci do not appear or appear with different alleles in NGS than in array-based genotyping. Here, four loci have been identified for which the use of alternative alleles could lead to the achievement of the intended, originally array-based determined AF, if NGS-based genotyping does not do so (Table 4). Two of these loci were absent in gnomAD v3.1.2 NFEs, which was also true for rs113778879 and rs572022984. As potential workaround for rs113778879, which is an SNV, an overlapping 5bp deletion was identified, but the observed AF exceeds the expected one by more than 0.1 (Supplementary Table 4). gnomAD SV v2.1 [23] reports a 1,370bp deletion starting at the same genomic position as rs572022984, namely DEL_2_27095, with an AF of 0.0417 in Europeans. However, genotyping of structural variants requires adapted variant calling approaches and therefore might be unfeasible within the scope of PRS genotyping in a routine diagnostic setting.

If no workarounds are available for loci showing noticeably deviating AFs, only imputation of the expected dosage according to CanRisk remains. This leads to smaller errors than omitting the locus from PRS calculation or setting the genotype to 0/0. However, each imputation causes a shift towards the mean PRS, and therefore imputations are meaningful only up to a certain extent.

PRSs for calculating individual BC risks will continue to evolve. For example, currently the Confluence Project^2^ aims to develop multi-ancestry PRSs. In addition, PRSs become also more and more relevant for diagnostics of other diseases with a genetic component [24,25]. The presented results underline that it would facilitate the implementation in clinical routine and thus also increase the reliability of genetic diagnostics if the design of future PRSs would be guided by the reproducibility of the expected AFs in addition to the observed effect sizes. A straightforward strategy to achieve this could be to ensure comparability of AFs in large-scale population databases, favorably based on different genotyping approaches, prior to including a locus in a PRS.

This study has limitations. Larger sample sizes may have resulted in more accurate estimators of AFs. Furthermore, there was a strong enrichment for samples derived from individuals with familial BC/OC, which may have resulted in deviating AFs due to genetic load rather than technical artifacts. The genetic background could explain, e.g., the aberrant (but concordant) AFs of rs851984 in ICG data and of rs35054928 in CFBOC data. Finally, no statement can be made about whether the described AF deviations would persist when using arrays for genotyping, since corresponding analyses are not (yet) performed in any of the GC-HBOC centers.

## Data Availability

All data generated or analyzed during this study are included in this published article [and its supplementary files].

## Acknowledgements

We thank the coordinator of the GC-HBOC, Rita K. Schmutzler, and all GC-HBOC center directors for their support of the GC-HBOC Bioinformatics Working Group. Further, we thank Joe Dennis for helpful comments.

**The Bioinformatics Working Group of the German Consortium for Hereditary Breast & Ovarian Cancer.** Norbert Arnold^13^, Alexandra Baumann^14^, Marvin Döbel^15^, Stephan Drukewitz^16^, Christoph Engel^17^, Corinna Ernst^18^, Rudel Christian Nkouamedjo Fankep^18^, Michael Forster^13^, Peter Frommolt^19^, Eva Groß^20^, Karl Hackmann^14^, Johannes Helmuth^21^, Ellen Honisch^22^, Tim Hutschenreiter^14^, Anna-Lena Katzke^23^, Anna-Lena Kobiela^18^, Zarah Kowalzyk^14^, Oliver Kutz^14^, Christoph Meier^24,25^, Maximilian Radtke^16^, Juliane Ramser^26^, Robert Remy^18^, Julia Ritter^21^, Christian Ruckert^27^, Gunnar Schmidt^23^, Benedikt Schnur^23^, Dariush Skowronek^28^, Marc Sturm^15^, Katharina Thiedig^26^, Steffen Uebe^29^, Shan Wang-Gohrke^30^, Andreas Zimmer^31^ ^13^Department of Gynecology and Obstetrics, Institute of Clinical Chemistry Institute of Clinical Molecular Biology, University Hospital Schleswig-Holstein, Campus Kiel, Kiel, Germany. ^14^Institute for Clinical Genetics, University Hospital Carl Gustav Carus at TU Dresden, Dresden, Germany; ERN GENTURIS, Hereditary Cancer Syndrome Center Dresden, Germany; National Center for Tumor Diseases Dresden (NCT/UCC), Germany: German Cancer Research Center (DKFZ), Heidelberg, Germany; Faculty of Medicine and University Hospital Carl Gustav Carus at TU Dresden, Dresden, Germany; German Cancer Consortium (DKTK), Dresden, Germany; German Cancer Re-search Center (DKFZ), Heidelberg, Germany; Max Planck Institute of Molecular Cell Biology and Genetics, Dresden, Germany. ^15^Institute of Medical Genetics and Applied Genomics, University Hospital Tübingen, Tübingen, Germany. ^16^Institute of Human Genetics, University of Leipzig Medical Center, Leipzig, Germany. ^17^Institute for Medical Informatics, Statistics and Epidemiology, University of Leipzig, Leipzig, Germany. ^18^Center for Familial Breast and Ovarian Cancer, Center for Integrated Oncology (CIO), Medical Faculty, University of Cologne and University Hospital Cologne, Cologne, Germany. ^19^Institute for Human Genetics, University Hospital Hamburg-Eppendorf, Hamburg, Germany. ^20^Department of Obstetrics and Gynecology, Ludwig-Maximilians-University of Munich, Munich, Germany. ^21^Department of Human Genetics, Labor Berlin – Charité Vivantes GmbH, Berlin, Germany. ^22^Department of Gynaecology and Obstetrics, University Hospital Düsseldorf, Heinrich-Heine University Düsseldorf, Düsseldorf, Germany. ^23^Department of Human Genetics, Hannover Medical School (MHH), Hannover, Germany. ^24^Institute of Human Genetics, University of Regensburg, Regensburg, Germany. ^25^Institute of Clinical Human Genetics, University Hospital Regensburg, Regensburg, Germany. ^26^Division of Gynaecology and Obstetrics, Klinikum rechts der Isar der Technischen Universität München, München, Germany. ^27^Institute of Human Genetics, University of Münster, Münster, Germany. ^28^Department of Human Genetics, University Medicine Greifswald and Interfaculty Institute of Genetics and Functional Genomics, University of Greifswald, Greifswald, Germany. ^29^Institute of Human Genetics, Universitätsklinikum Erlangen, Friedrich-Alexander-Universität, Erlangen-Nürnberg, Germany. ^30^Department of Gynaecology and Obstetrics, University Hospital Ulm, Ulm, Germany. ^31^Institute for Human Genetics, Medical Center University of Freiburg, Faculty of Medicine, University of Freiburg, Freiburg, Germany.

## Author contributions

Conceptualization: GS, MS and CE. Methodology: All authors. Data analysis: AB, CR, CM, RR, MS and CE. Editing and review of manuscript: All authors. Final manuscript review: All authors.

## Ethical approval

IMGAG: The use of aggregate statistics of human subject genetics data was approved by the ethics committee of the Medical Faculty of the University of Tübingen, Germany (Genome+, ClinicalTrial.gov-Nr: NCT04315727; #066/2021BO2 for retrospective data analysis). ICG, IHG-M, CFBOC, IHG-R: Written informed consent was obtained from all patients and ethical approval was granted by the ethics committee of the Technische Universität Dresden, ethics committee of the Medical Association Westfalen-Lippe, ethics committee of the Medical Faculty of the University of Cologne (19-1360_4), the ethics committee of the University of Regensburg (21-2192-103).

## Footnotes

1 https://gnomad.broadinstitute.org

2 https://confluence.cancer.gov

