## Supplementary Material for "Limitations in next-generation sequencing-based genotyping of breast cancer polygenic risk score loci"

Alexandra Baumann, Christian Ruckert, Christoph Meier, Tim Hutschenreiter, Robert Remy, Benedikt Schnur, Marvin Döbel, Rudel Christian Nkouamedjo Fankep, Dariush Skowronek, Oliver Kutz, Norbert Arnold, Anna-Lena Katzke, Michael Forster, Anna-Lena Kobiela, Katharina Thiedig, Andreas Zimmer, Julia Ritter, Bernhard H.F. Weber Ellen Honisch, Karl Hackmann, Bioinformatics Working Group of the German Consortium for Hereditary Breast & Ovarian Cancer, Gunnar Schmidt, Marc Sturm, Corinna Ernst

### Supplementary Methods

#### Identification of alternative alleles and proxys

The gnomAD v3.1.2 web interface was employed to search for overlapping alternative variants located within 20bp of each polygenic risk score (PRS) locus showing noticeably deviating allele frequency (AF) in at least one real-world setting under consideration.

For the identification of proxys, i.e., single-nucleotide variants (SNVs) or indels in linkage disequilibrium, the LDproxy utility of the LDlink web interface [4] and TopLD [3] were employed. LDproxy utilizes data from the 1000 Genomes Project (1000G) to compute correlations and AFs. Proxys for PRS loci were retrieved based on the microarray-genotyped GRCh37 [9] and GRCh38 High Coverage WGS data [1] of the Utah Residents from North and West Europe (CEU) population.

TopLD results are based on WGS data of the European population from the NHLBI Trans-Omics for Precision Medicine (TOPMed) program [8]. Both for LDproxy and TopLD, loci were passed via their dbSNP identifiers [7] and then the optimal proxy was determined by first maximum $R^2$ and second minimum distance in base pairs. As in case of adjacent proxys genotyping might be accompanied by similar technical difficulties as for the original locus, each the next closest non-adjacent variant was taken into account (e.g., as substitutes for locus rs9421410, rs7913694 and rs35098964 were considered instead of rs9421409). Only loci in linkage disequilibrium with a minimum value of $R^2 = 0.8$ were reported. Proxy pairs retrieved by LDproxy, were checked using the LDpair utility of LDlink, and only those combinations were reported, that appeared accordingly in LDpair, i.e., the alleles of the reference locus matched the CanRisk expectations and the AFs of the proxy matched the AF formerly reported by LDproxy.

#### Real-world data generation

The Insititute of Medical Genetics and Applied Genomics (IMGAG), University Hospital Tübingen, provided results of BCAC 313 BC PRS genotyping based on WGS of 1758 individuals who underwent genetic testing due to various indications, i.e., not exclusively due to suspicion of hereditary BC/OC. Data was obtained based on hg38 reference, using the megSAP pipeline[1], including genotyping of a subset of 348 samples with DRAGEN v4.0.3 [5] and of 1410 samples with freebayes v1.3.6 [2]. Both variant callers were run in unforced cohort mode, i.e., no lists of variants to be genotyped were

[1] https://github.com/imgag/megSAP/

passed, and only calls reported with a minimum sequencing coverage of 15 were considered in AF computation.

The Institute for Clinical Genetics (ICG), University Hospital Carl Gustav Carus Dresden, provided data of overall 595 European individuals, of whom 372 underwent genetic testing due to familial BC/OC and 223 due to different (cancer-related) indications. Sequencing was done using a Custom Cancer Panel (Twist Bioscience). Reads were mapped to the hg19 reference and forced genoyping was applied via specification of `--variant-input` with freebayes v1.3.6 and `--call` with GATK v4.2.6 HaplotypeCaller [6] for forced genotyping of the overall 324 loci of both the BCAC 313 and the BRIDGES 306 BC PRS. Loci with read depth $<20$ in corresponding VCF outputs were excluded from AF computation.

The Institute of Human Genetics at the University of Münster (IHG-M) provided results of hg19-based BRIDGES 306 BC PRS genotyping of 593 European individuals who have underwent genetic testing due to familial BC/OC. Sequencing was performed using a Twist Custom Panel, covering exonic regions of 130 genes and selected variants. Genotyping was performed using DRAGEN v4.2.4 and GATK HaplotypeCaller v4.4.0, each in forced genotyping mode. Only loci with read depth $\geq 20$ in corresponding VCF outputs were considered for AF computation.

The Center for Familial Breast an Ovarian Cancer (CFBOC), University Hospital Cologne, provided genotyping results of 416 samples from European individuals with BC/OC family history, based on TruRisk™ v3 hybridization-based capture sequencing (Agilent SureSelect, QXT protocol), using the hg19 reference and forced genotyping with freebayes v1.3.6 [2] and GATK v4.3.2 HaplotypeCaller. The loci of the BRIDGES 306 BC PRS were considered, and only those with a minimum read depth of 30 in corresponding VCF outputs were included in AF computation.

The Institute of Human Genetics at the University of Regensburg (IHG-R) provided results of BRDIGES 306 BC PRS genotyping of 251 European individuals who have underwent genetic testing due to familial BC/OC. Sequencing was done using Agilent TruRisk-Panel™ v3. Genotyping was performed using both the CLC LightSpeed Module 23.0.2 from CLC Genomics Workbench 23.0.3, which provides an automated workflow from read trimming up to variant calling, and an inhouse workflow using publicly available standard tools for NGS analysis, including GATK HaplotypeCaller v4.2.6.1 for forced genotyping.

### Supplementary Tables

Supplementary Table 1: Observed allele frequencies (AFs) of 325 BCAC 313 or BRIDGES 306 breast cancer polygenic risk score (PRS) loci in gnomAD v3.12 non-Finnish Europeans and technical artifacts reported.

Supplementary Table 2: Polygenic risk score (PRS) genotyping results in ten real-world settings. Log odds ratios (ORs) refer to effect sizes. Data was provided by the Insititute of Medical Genetics and Applied Genomics (IMGAG) at University Hospital Tübingen, Institute for Clinical Genetics (ICG) at University Hospital Carl Gustav Carus Dresden, by the Institute of Human Genetics at the University of Münster (IHG-M), by the Center for Familial Breast and Ovarian Cancer (CFBOC) at University Hospital Cologne, and by the Institute of Human Genetics at the University of Regensburg (IHG-R). AF = allele frequency.

Supplementary Table 3: Estimated 10 year (e10yr) and remaining lifetime risks (eLTR) of developing primary breast cancer for cancer-unaffected women aged 20, 40, and 60 years due to CanRisk, including artifical polygenic risk scores, constructed by subtitution of expected dosages by observed noticeably deviating allele frequenices in real-world settings.

Supplementary Table 4: Overlapping alternative variants located within 20bp of each polygenic risk score locus showing noticeably deviating allele frequencies (AFs) in real-world data (listed in Table 2). Only alternative variants with AF$\geq$0.01 in gnomAD v3.1.2 non-Finnish Europeans are listed.

Supplementary Table 5: Loci in linkage disequilibrium (proxys) to 24 polygenic risk score loci showing noticeably deviating allele frequencies (AFs) in real-world data (listed in Table 2). Only loci with $R^2 \geq 0.8$ due to LDproxy or TopLD are listed. MAF: Minor AF.

### Supplementary Figures

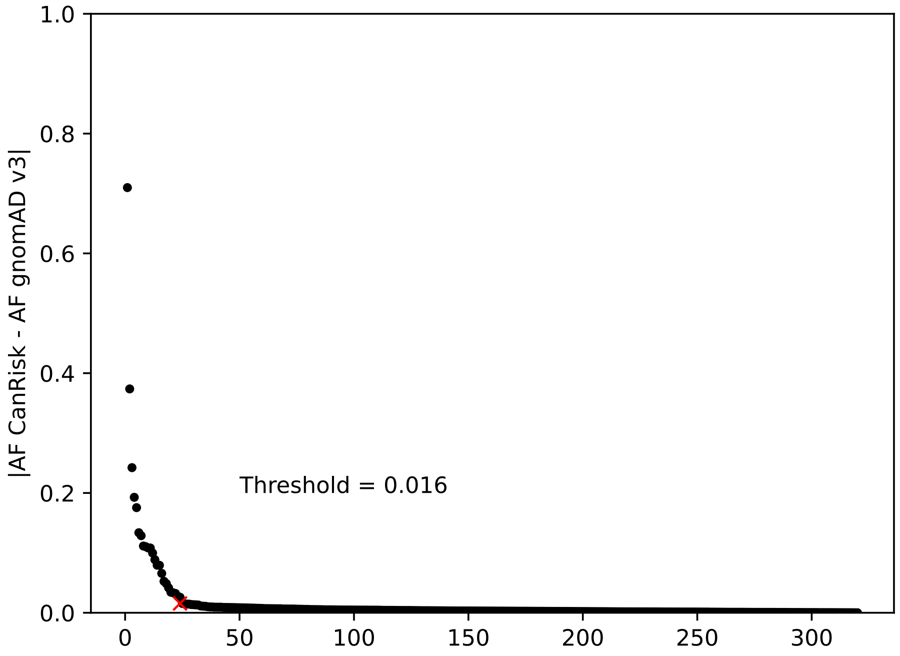


Supplementary Figure 1: Elbow in the curve analysis of absolute differences between expected and observed allele frequencies in gnomAD v3.1.2 of the BCAC 313 breast cancer (BC) and BRIDGES 306 BC polygenic risk score loci.

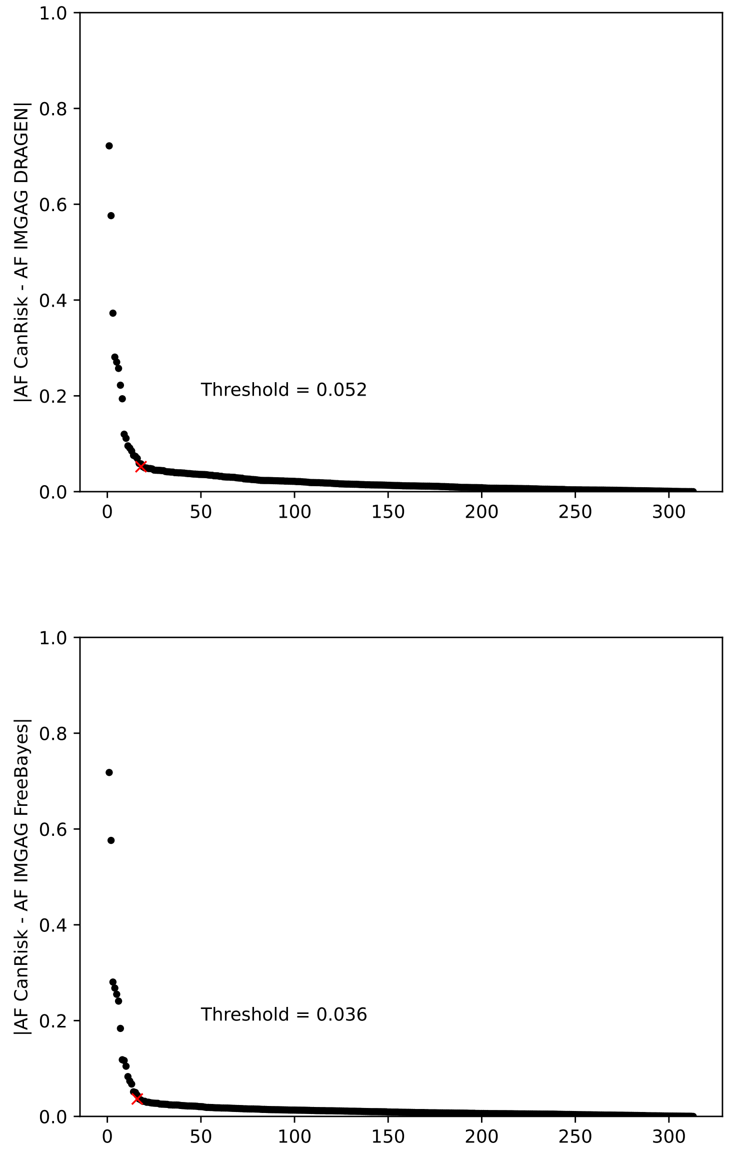


Supplementary Figure 2: Elbow in the curve analysis of absolute differences between expected and observed allele frequencies (AFs) of BCAC 313 breast cancer (BC) polygenic risk score loci in DRAGEN-derived data based on 348 samples (above) and freebayes-derived data based on 1410 samples (below). Data was provided by the Institute of Medical Genetics and Applied Genomics (IMGAG) at University Hospital Tübingen.

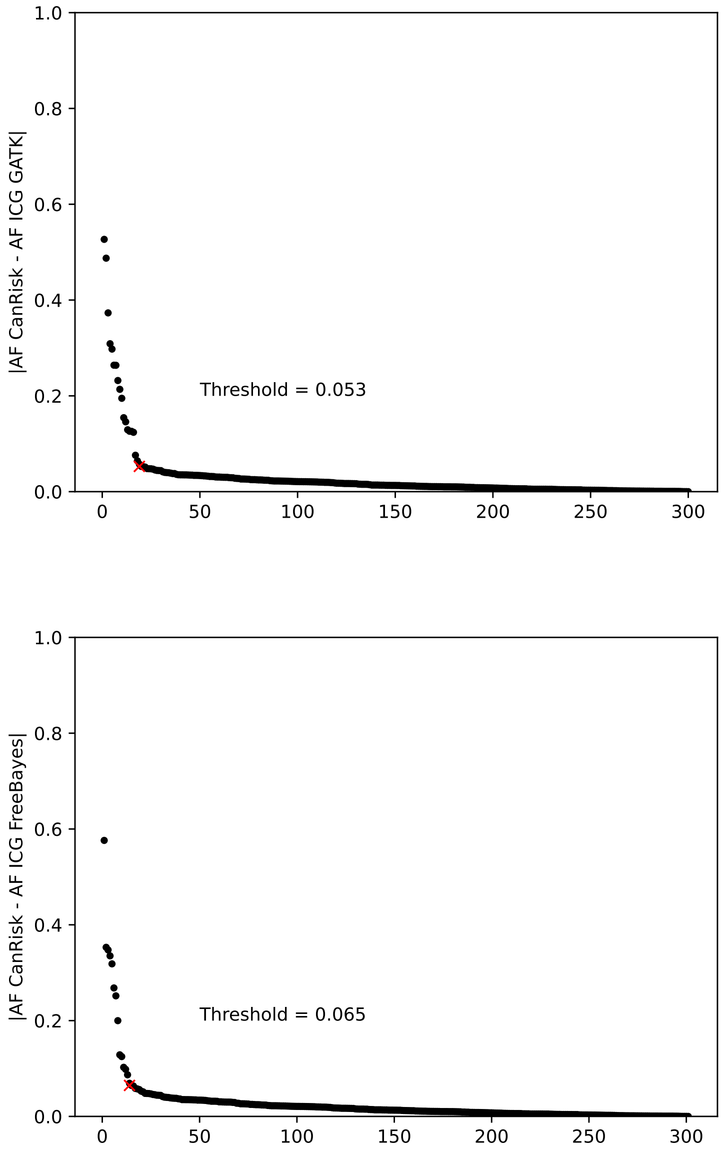


Supplementary Figure 3: Elbow in the curve analysis of absolute differences between expected and observed allele frequencies (AFs) of BCAC 313 breast cancer (BC) and BRIDGES 306 BC polygenic risk score loci in GATK-derived (above) and freebayes-derived (below) data based on 595 samples. Data was provided by the Institute of Clinical Genetics (ICG) at University Hospital Carl Gustav Carus Dresden.

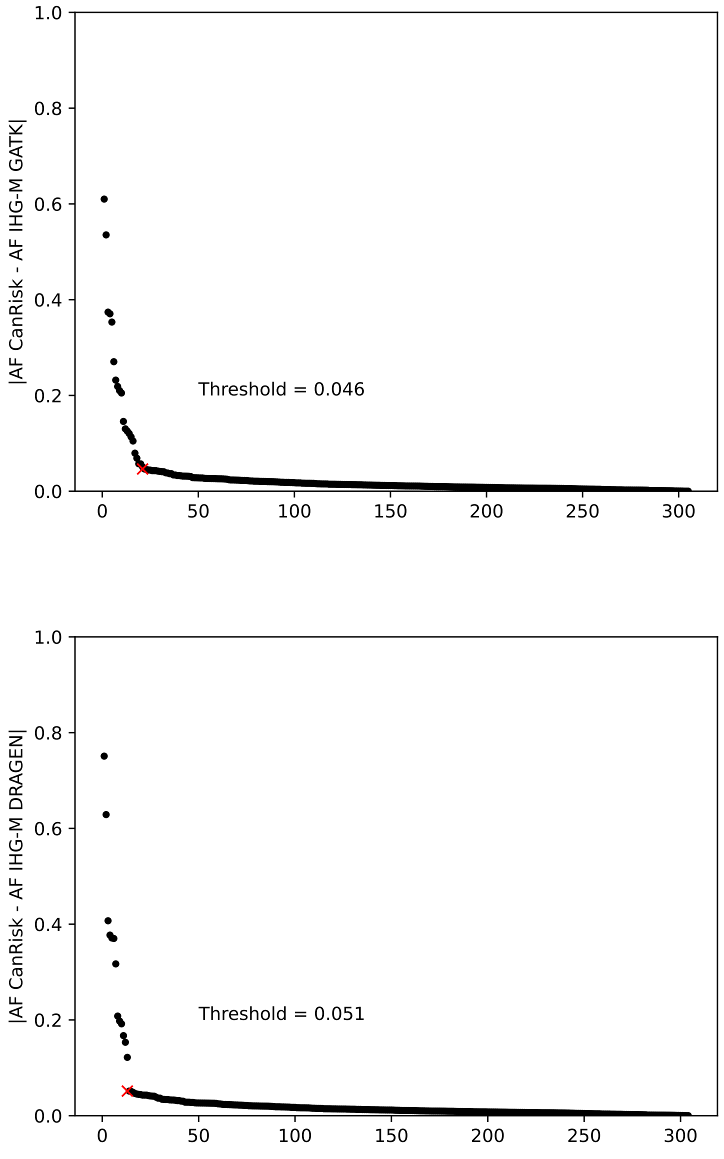


Supplementary Figure 4: Elbow in the curve analysis of absolute differences between expected and observed allele frequencies (AFs) of BRIDGES 306 breast cancer polygenic risk score loci in GATK-derived (above) and DRAGEN-derived (below) data based on 593 samples. Data was provided by the Institute of Human Genetics at the University of Münster (IHG-M).

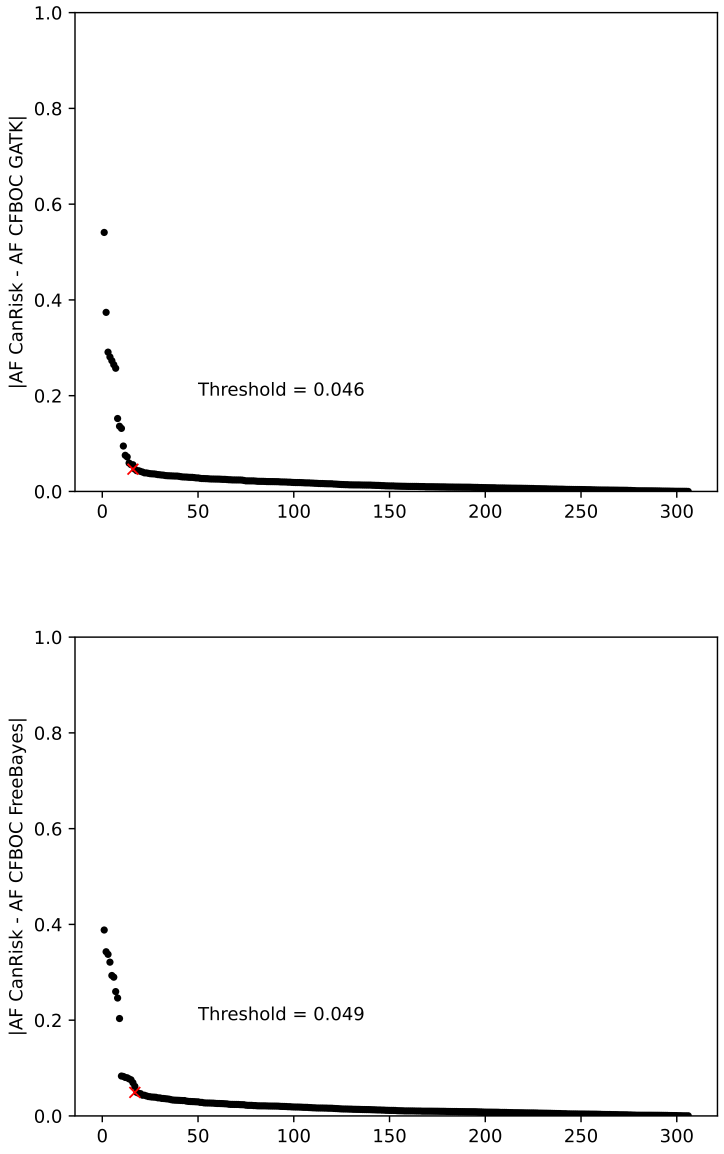


Supplementary Figure 5: Elbow in the curve analysis of absolute differences between expected and observed allele frequencies (AFs) of BRIDGES 306 breast cancer polygenic risk score loci in GATK-derived (above) and freebayes-derived (below) data based on 416 samples. Data was provided by the Center for Familial Breast and Ovarian Cancer (CFBOC) at University Hospital Cologne.

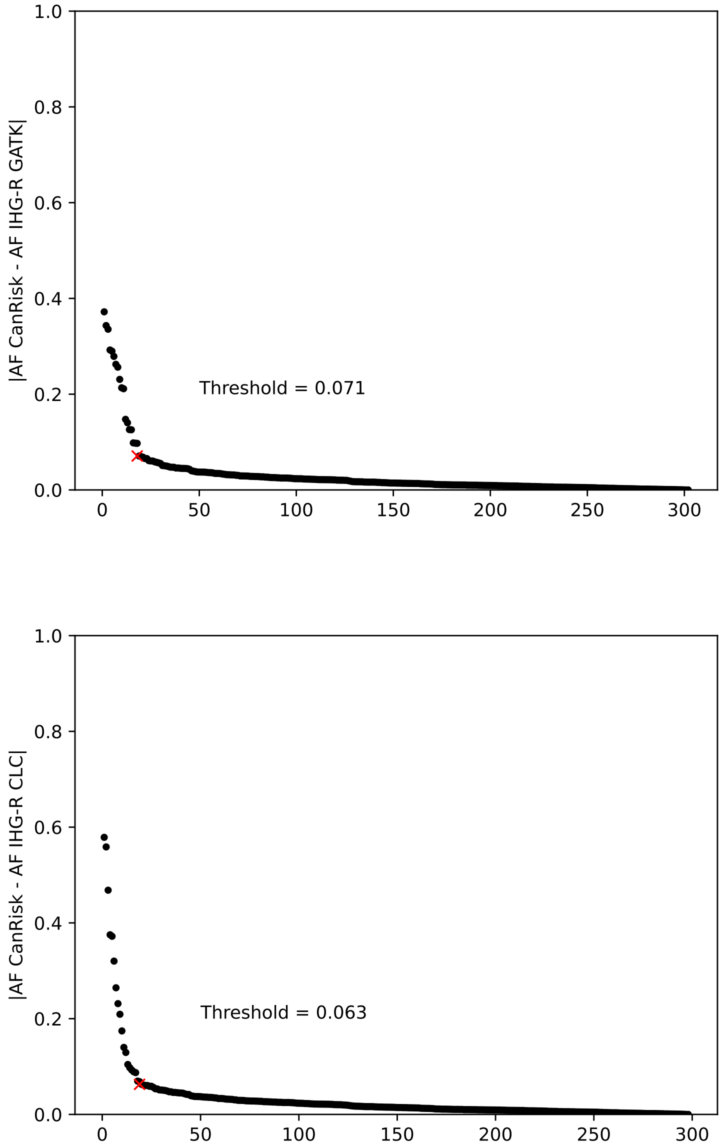


Supplementary Figure 6: Elbow in the curve analysis of absolute differences between expected and observed allele frequencies (AFs) of BRIDGES 306 breast cancer polygenic risk score loci in GATK-derived (above) and CLC LightSpeed Module-derived (below) data based on 251 samples. Data was provided by the Institute of Human Genetics at the University of Regensburg (IHG-R).

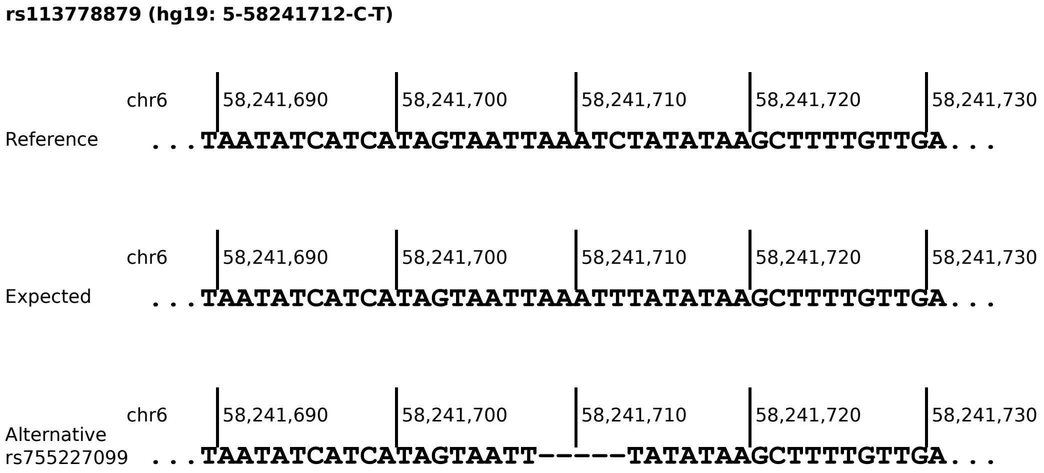


Supplementary Figure 7: Sequences of reference, expected effect allele and potential alterantive allele of polygenic risk locus rs113778879 (hg19-based).
